# ARE NUTRITIONAL ASPECTS AND BODY COMPOSITION ASSOCIATED WITH THE “CAN DO, DO DO” CONCEPT IN PEOPLE WITH COPD IN LATIN AMERICA? AN OBSERVATIONAL STUDY

**DOI:** 10.64898/2026.04.13.26350788

**Authors:** Paloma Borges, Ana Paula Coelho Figueira Freire, Matheus Pedroso, Bruna Spolador de Alencar Silva, Fabiano Francisco de Lima, Juliana Souza Uzeloto, Luis Alberto Gobbo, Isis Grigoletto, Ercy Mara Cipulo Ramos

**Author notes:** **Corresponding author:** Paloma Borges., Department of Physiotherapy, São Paulo State University (UNESP), School of Technology and Science., Roberto Simonsen Street, 305, Centro Educacional, Zip-Code: 19060-900, Presidente Prudente, São Paulo, Brazil.

## Abstract

**Introduction:** Individuals with COPD can be classified according to their levels of physical activity (PA) and physical capacity (PC). The relationship between nutrition and body composition within these classifications remains unclear.

**Objectives:** To compare the body composition and food intake of people with COPD and verify the associations.

**Methods:** Cross-sectional exploratory analysis study in which body composition and food intake were assessed in individuals with COPD. Classification was based on six-minute walk test (PC) and accelerometry(PA): Quadrant “can do, don’t do” (I-preserved PC, low PA); quadrant “can do, do do” (II-preserved PC, preserved PA).

**Results:** 72 individuals with COPD, 39 in quadrant I and 33 in quadrant II, with mean ages of (69 ± 6) (67 ± 7), respectively. Group I had a higher proportion of males, whereas group II had a higher proportion of females. A positive trend in skeletal muscle mass (p=0.011) (B= 2.883) and a negative trend in basal metabolic rate (p=0.010) (B=-0.092) for group I.

**Conclusion:** Brazilians with COPD classified in quadrants I and II showed similar results in terms of body composition and food intake. A positive trend in skeletal muscle mass was observed for the group I. These findings align with the pathophysiological model of COPD, in which the preservation of muscle mass and adequate protein intake support functional capacity and the maintenance of higher physical activity levels.

## 1. BACKGROUND

Both physical capacity (PC) and physical activity (PA) levels are associated with better prognoses in patients with COPD. However, individuals who are physically capable of exercising do not always engage in such activities^1-6^. PA level is an important prognostic factor for hospitalization and mortality, and it can be negatively affected by the side effects of pharmacological therapy ^1, 7, 8^ . Physical capacity increases directly with PA and can be improved through exercise training^9^.

Given this context, Koolen et al. (2019) developed a classification system based on PA and PC for individuals with COPD, with the aim of investigating the relationship between PA, PC, and clinical characteristics. They demonstrated that this classification may serve as a useful tool for understanding physical function impairment in individuals with COPD^10^. In their study, the authors recommended that “to address behavioral change at the individual level, all personal barriers and facilitators to PA engagement should be considered in future studies” ^10^.

Considering that COPD is commonly associated with malnutrition, driven by increased metabolic demands due to respiratory effort, it is reasonable to assume that nutrition and body composition may influence the classification of individuals within the proposed quadrants^7, 11-15^. These factors contribute to alterations in body composition, which represents one of the modifiable parameters in this population. In addition, several other factors may lead to loss of lean mass and an increase in fat mass, such as corticosteroid use, hormonal changes, physical inactivity, and increased energy expenditure resulting from inflammation and hypoxia ^10, 15, 16^. Given that reductions in lean mass impair muscle strength and endurance—both of which affect PA, a key determinant of functional capacity—understanding these relationships is essential^17,18^.

Nutritional deficiency can negatively affect exercise performance, muscle function, and lung function, and is also associated with increased exacerbations, higher mortality, and greater healthcare costs^10, 11, 19^. Therefore, nutritional aspects are highly relevant for individuals with COPD and may act as either facilitators or barriers to PA. In the context of poor nutrition, sarcopenia has an even greater impact in this population, further limiting PA and reducing PC^20^.

Understanding how nutritional aspects and body composition interact with PA and PC in individuals with COPD may help identify opportunities for targeted nutritional and exercise interventions. Such adjustments in those classified within the “don’t do” quadrants could significantly improve PA levels and potentially enhance quality of life by reducing exacerbations and symptoms.

The objective of this study was to compare the body composition and food intake of individuals diagnosed with COPD using data from a secondary study with an exploratory analysis of the “Can do, don’t do” (adequate PC and low PA) and “Can do, do do” (adequate PC and adequate PA) quadrants proposed by Koolen et al. (2019). In addition, the study aimed to examine the associations between these outcomes and the quadrants characterized by the “can do” ability.

## 2. METHODS

### 2.1. Study design

This cross-sectional observational study involved an exploratory analysis of secondary data from a study conducted in Presidente Prudente, Brazil, in which individuals with COPD were invited to participate. The study was approved by the local Research Ethics Committee (in accordance with the Declaration of Helsinki; protocol 77909317.2.0000.5402), and written informed consent was obtained from all participants.

Assessments included body composition (octopolar bioelectrical impedance), physical capacity (6MWT), dietary intake (three-day food record), and physical activity (accelerometry).

The inclusion criteria were: (1) a diagnosis of COPD according to the Global Initiative for Chronic Obstructive Lung Disease (GOLD)^14^; (2) clinical stability, with no exacerbations or changes in medication for at least 30 days; (3) no use of home oxygen therapy; and (4) absence of pathological conditions that could limit PA. The exclusion criteria were: (1) incomplete nutritional records; and (2) failure to complete the tests required for group classification (6MWT and accelerometry).

The sample size for this secondary analysis was determined using the mean and standard deviation of the total distance covered in the 6MWT, given that physical capacity is one of the primary outcomes for comparing the two groups. Considering a 20% sample loss, a total of 72 participants was estimated to provide 80% power with a 95% confidence interval and a significance level of 5%^10^.

### 2.2. Assessments

#### 2.2.1. Initial assessment

These individuals were firts interview and assessed to collect personal and anthropometric data, as well as to confirm the diagnosis using a lung function test, in accordance to the standards of the American Thoracic Society (ATS) e European Respiratory Society (ERS)^21^, with reference values for the Brazilian population^22^. The diagnosis of COPD was confirmed using a portable spirometer MIR–Spirobank version 3.6.

#### 2.2.2. Body composition

Body composition was assessed using the InBody 720 octopolar bioelectrical impedance device (Biospace, Seoul, Korea), which provides variables such as weight, fat mass, visceral adiposity area, fat-free mass (FFM), skeletal muscle mass (SMM), lean body mass (LBM), total body water (TBW), protein content, and mineral content. The data were electronically imported into Excel using the Lookin’Body 3.0 software (Biospace, Seoul, Korea).

#### 2.2.3. Assessment of nutritional intake

Nutritional intake was assessed using a food diary. Participants were instructed to record their food consumption over three days (on nonconsecutive days, including at least one weekend day), as this is the minimum period required for a quantitative assessment of nutrient intake and food group consumption. Portion sizes and preparation methods (or the brand name, in the case of processed foods) were recorded in detail. To improve the accuracy of portion size estimation, participants were instructed to use standard household measures (e.g., teaspoons, tablespoons, and cups) and to take photographs of the different portion sizes. ^23, 24^

#### 2.2.4. Physical Capacity assessment

To analyze physical capacity, the Six Minute Walk Test (6MWT) was performed according to the guidelines established by the European Respiratory Society and the American Thoracic Society^25, 26^. In this test, the individual needs to walk a distance of 30 meters for six continuous minutes, and the distance covered will be calculated.

#### 2.2.5. Physical Activity assessment

To analyze habitual PC, an Actigraph accelerometer-type motion sensor, model GT3X (Actigraph LLC, Pensacola, FL). This device was used for seven days and PA was expressed as the average number of steps per day measured during at least four valid days ^27^.

#### 2.2.6. Classification of subgroups

The classification proposed by Koolen et al. involved four quadrants based on the number of steps per day, measured by an accelerometer, and the percentage of predicted results from (6MWT)^10^, as in figure 1. Koolen et al classified quadrant I as to include individuals with low levels of PC (6MWT <70% of the predicted value), indicating they “Can’t do” and PA (<5.000 steps/day), indicating they “don’t do”. Quadrant II: preserved PC (6MWT >70% of the predicted value), indicating individuals that “Can do” and low PA (<5.000 steps/day) however they “don’t do”. Quadrant III: low PC (6MWT <70% to predicted), indicating they “Can’t, do” and preserved PA (>5.000 steps/day) indicating they “do do” and Quadrant IV: preserved PC (6MWT >70% of the predicted value) indicating they “Can do” and preserved PA (>5.000 steps/day) indicating they “do do”^10^.

**Figure 1.**
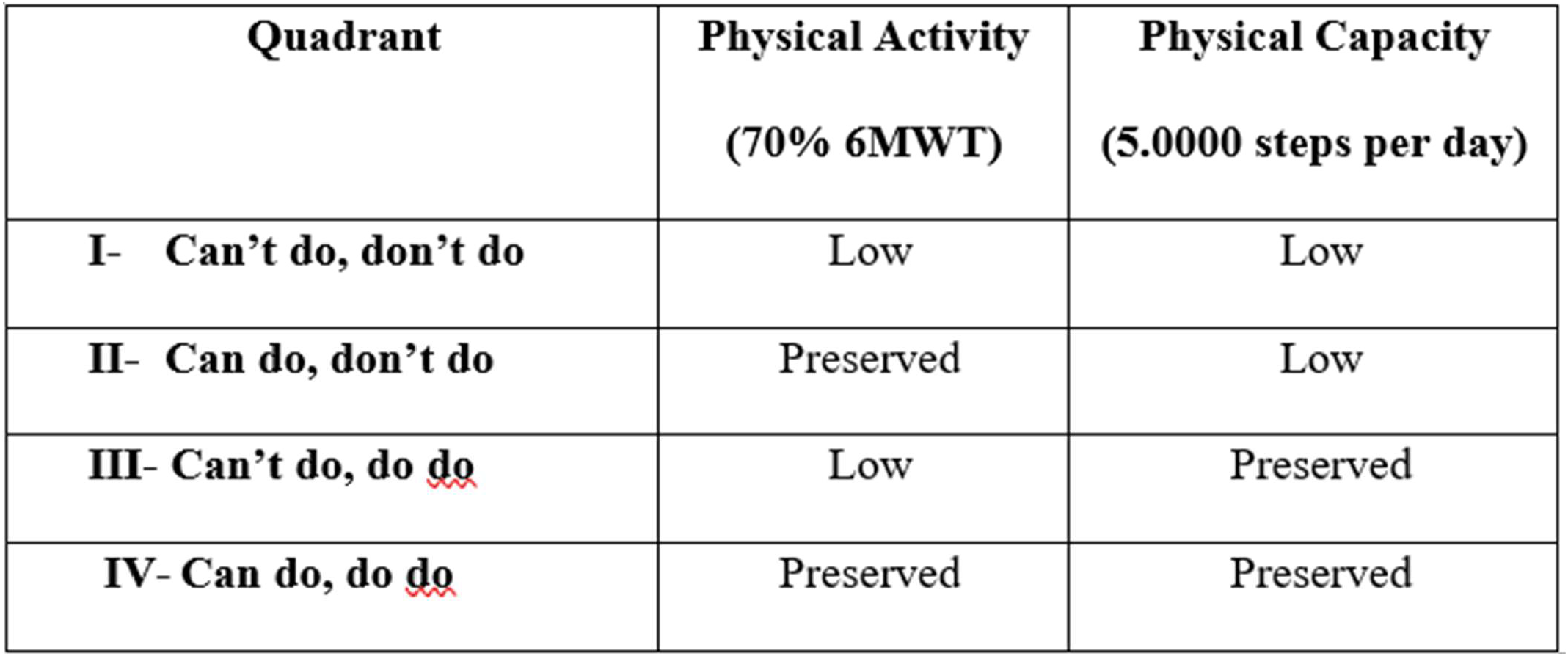
Distribution of quadrants

### 2.3. Statistical methods

For statistical analysis, the Statistical Package for the Social Sciences (SPSS) version 29.0 was used. Data were subjected to the Kolmogorov–Smirnov test to assess normality. Variables with a normal distribution were expressed as mean and standard deviation, whereas non-normally distributed variables were presented as median and interquartile range. The significance level was set at 5%.

Group comparisons were performed using the Mann–Whitney test for non-parametric data and Student’s t-test for parametric data. Binary logistic regression models were applied to evaluate the odds and outcomes of the dependent variable (classification into the “Can do, do do” or “Can do, don’t do” groups) in relation to nutritional intake and body composition variables considered as independent predictors.

The selection of variables for logistic regression was based on their potential relevance to PA, such as skeletal muscle mass (SMM), protein intake, and lean body mass (LBM).

## 3. RESULTS

Seventy-nine individuals were recruited, of whom 72 were included in the analysis of this secondary study. Participants were classified into the “Can do, does not do” group (n = 39) or the “Can do, does do” group (n = 33). Due to the small number of participants in the remaining quadrants, the “Can do, does” and “Can’t do, does” groups were not included in the comparative statistical analyses. The study flowchart is presented in Figure 2.

**Figure 2.**
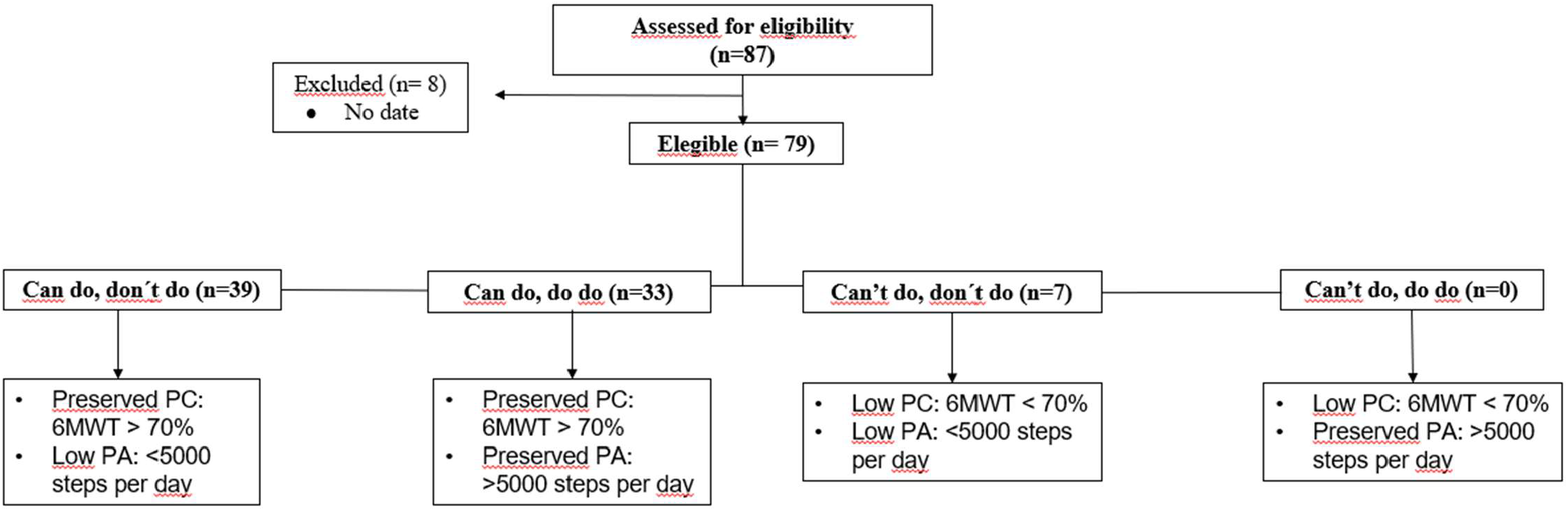
Flowchat

Table 1 provides the characterization of the sample, including lung function, functional capacity, dyspnea, physical activity, nutritional status, and body composition measured by bioelectrical impedance (BIA), stratified by group.

**Table 1.** Comparison between the quadrants proposed by Koolen et al.

| Sample Size (n=72) |  |  |  |
| --- | --- | --- | --- |
|  | “Can do, don’t do” (n=39) | “Cand do, do do” (n=33) | p-value |
| Sex (M/F) | 22/17 | 12/21 | 0,090 |
| Age (Years) | 69 ± 6 | 67 ± 7 | 0,208 |
| BMI (kg/cm<sup>2</sup>) | 27 ± 5 | 26 ± 5 | 0,523 |
| Rehabilitation (n/%) | 9/23 | 9/27 | 0,682 |
| **Lung function** |  |  |  |
| FVC (%pred) | 69 ± 16 | 77 ± 11 | 0,443 |
| FEV<sub>1</sub> (%pred) | 53 ± 17 | 54 ± 14 | 0,661 |
| FEV<sub>1</sub>/FVC (%) | 55 ± 12 | 56 ± 11 | 0,805 |
| **Functional capacity** |  |  |  |
| 6MWT(pred) | 530 ± 35 | 527 ± 39 | 0,739 |
| 6MWT (m) | 470 ± 63 | 515 ± 72 | 0,006* |
| **Acelerometry (minutes)** |  |  |  |
| Step per day | 3572 [2747 – 4278] | 6096 [5387 – 8601] | <0,001* |
| Sedentary time | 1015 ± 165 | 1069 ± 130 | 0,121 |
| **Nutritional intake** |  |  |  |
| Total food intake (g/d) | 1061 ± 368 | 1178 ± 545 | 0,303 |
| Caloric intake (Kcal/d) | 1183 [973 – 1513] | 1384 [976 – 1663] | 0,296 |
| Protein intake (g/dl) | 63 [48 – 70] | 66 [46 – 99] | 0,527 |
| Carbohydrate intake (d)g | 143 ± 54 | 152 ± 64 | 0,522 |
| **Body composition** |  |  |  |
| Basal metabolic rate (Kcal) | 1359 ± 176 | 1277 ± 158 | 0,040* |
| Visceral fat area (cm<sup>2</sup>) | 114 ± 37 | 10 ± 36 | 0,258 |
| Skeletal muscle mass (kg) | 25 ± 5 | 23 ± 5 | 0,066 |
| Body fat mass (kg) | 27 ± 10 | 25 ± 10 | 0,492 |
| Total body water (l) | 34 ± 6 | 31 ± 5 | 0,038* |
| Protein mass (Kg) | 9 [8 – 10] | 9 [7 – 9] | 0,082 |
| Mineral mass (Kg) | 3 [3 – 4] | 3 [3 – 3] | 0,058 |
| Fat free mass (Kg) | 46 ± 8 | 42 ± 7 | 0,039* |
| Lean body mass (Kg) | 43 ± 8 | 40 ± 7 | 0,044* |

BMI: body mass index; kg: kilograms; cm<sup>2</sup>: square centimeter; n: number; %: percentage; FVC: forced vital capacity; FEV<sub>1</sub>: forced expiratory volume in the first second; pred: predict; 6MWT:six-minute walk test; g: grams; d: day; kcal: kilocalories; L: liters. \*: p<0.05. Data expressed as mean and standard deviation or median and interquartile range 25-75%

### 3.1 Dietary Intake

Food intake parameters did not show statistically significant differences between the two groups analyzed. The p-values were as follows: total food intake (p = 0.303), caloric intake (p = 0.296), protein intake (p = 0.527), and carbohydrate intake (p = 0.522). These results are presented in Table 1.

### 3.2 Body Composition

Body composition values were higher in the “Can do, don’t do” group compared to the “Can do, do do” group, with statistically significant differences in basal metabolic rate (BMR) (p = 0.040), total body water (TBW) (p = 0.038), fat-free mass (FFM) (p = 0.039), and lean body mass (LBM) (p = 0.044). These data are shown in Table 1.

### 3.3 Associations between body composition, nutritonal intake and quadrants

Logistic regression analyses were performed using bioimpedance and food intake variables. Skeletal muscle mass emerged as a strong positive predictor (B = 2.883; p = 0.011) of belonging to the functionally active group (“Can do, do do”). Conversely, basal metabolic rate showed a negative association with this group (B = –0.092; p = 0.010) (Table 2). Additionally, an analysis combining muscle-related bioimpedance variables with protein intake—key components for physical capacity and activity—did not reveal significant associations (Table 3).

**Table 2.**
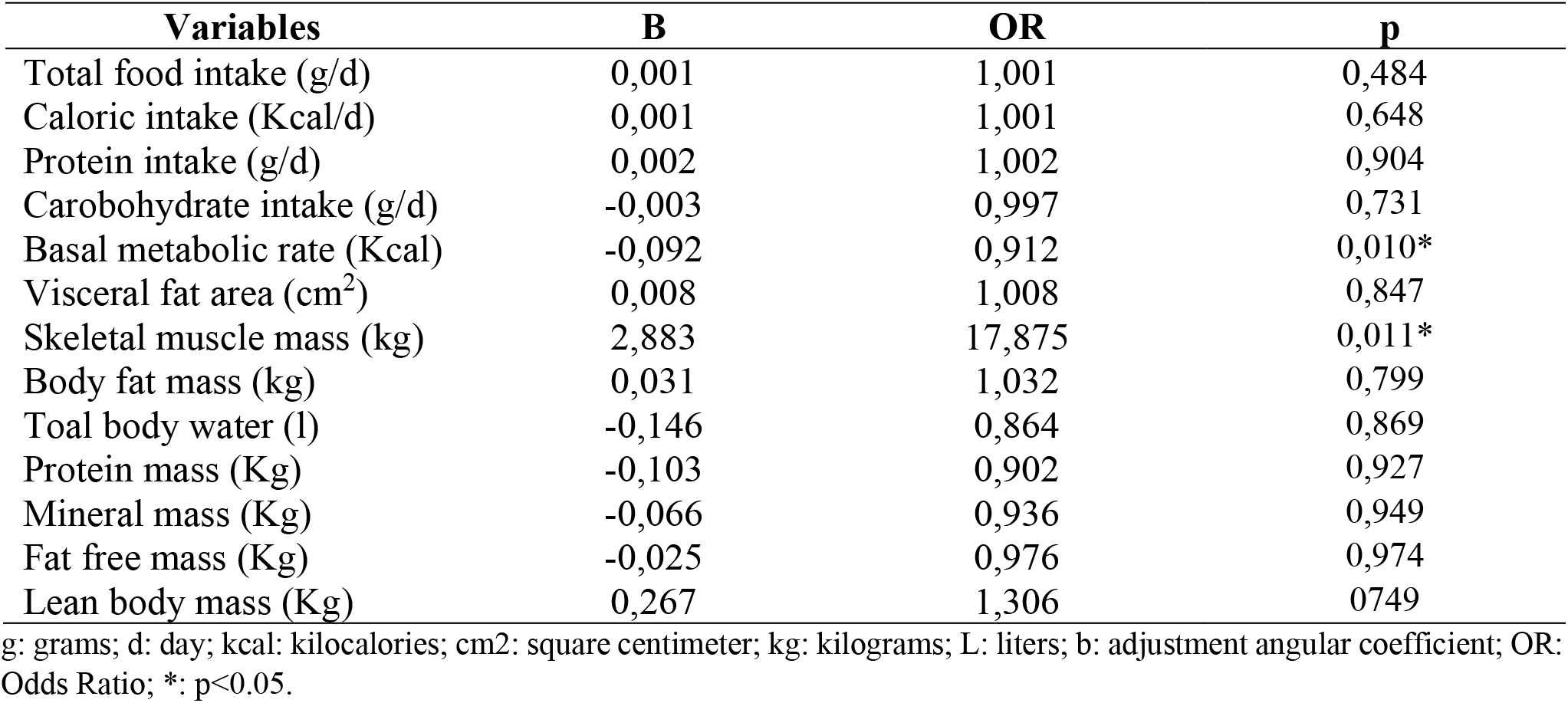
Simple logistic regression results. Associations between body composition and nutritional intake for the group “can do, do do”.

| Variables | B | OR | p |
| --- | --- | --- | --- |
| Total food intake (g/d) | 0,001 | 1,001 | 0,484 |
| Caloric intake (Kcal/d) | 0,001 | 1,001 | 0,648 |
| Protein intake (g/d) | 0,002 | 1,002 | 0,904 |
| Carbohydrate intake (g/d) | -0,003 | 0,997 | 0,731 |
| Basal metabolic rate (Kcal) | -0,092 | 0,912 | 0,010* |
| Visceral fat area (cm<sup>2</sup>) | 0,008 | 1,008 | 0,847 |
| Skeletal muscle mass (kg) | 2,883 | 17,875 | 0,011* |
| Body fat mass (kg) | 0,031 | 1,032 | 0,799 |
| Total body water (l) | -0,146 | 0,864 | 0,869 |
| Protein mass (Kg) | -0,103 | 0,902 | 0,927 |
| Mineral mass (Kg) | -0,066 | 0,936 | 0,949 |
| Fat free mass (Kg) | -0,025 | 0,976 | 0,974 |
| Lean body mass (Kg) | 0,267 | 1,306 | 0,749 |

g: grams; d: day; kcal: kilocalories; cm<sup>2</sup>: square centimeter; kg: kilograms; L: liters; b: adjustment angular coefficient; OR: Odds Ratio; \*: $p < 0.05$ .

**Table 3.** Simple logistic regression results. Associations between some variables important for de body for the group “can do, do do”.

| Variables | B | OR | p |
| --- | --- | --- | --- |
| Protein intake (g/d) | 0,012 | 1,012 | 0,155 |
| Skeletal muscle mass (kg) | 0,355 | 1,426 | 0,429 |
| Protein mass (Kg) | 0,492 | 1,636 | 0,594 |
| Fat free mass (Kg) | -0,581 | 0,559 | 0,321 |
| Lean body mass (Kg) | 0,203 | 1,225 | 0,759 |

g: grams; d: day; kcal: kilocalories; cm<sup>2</sup>: square centimeter; kg: kilograms; L: liters; b: adjustment angular coefficient; OR: Odds Ratio; \*: $p < 0.05$ .

Adjustments for sex and alternative combinations of bioimpedance and food intake variables were performed, but no statistically significant or clinically relevant differences were identified for further discussion.

## DISCUSSION

Our study included participants with COPD classified into the “Can do, don’t do” and “Can do, do do” quadrants proposed by Koolen et al. We identified that both groups had similar nutritional intake, while the “Can do, don’t do” group presented higher values in several bioimpedance variables. Although these values were within the normal range, statistically significant differences were observed in basal metabolic rate, total body water, fat-free mass, and lean body mass.^28, 29^

According to the logistic regression results, skeletal muscle mass was the main positive predictor of belonging to the “Can do, do do” group, as defined by Koolen et al. This indicates that individuals with COPD who have greater skeletal muscle mass are more likely to maintain preserved functional capacity and adequate levels of PA. This finding aligns with the literature demonstrating a direct relationship between muscle mass and functional performance in COPD^30, 31^. Loss of lean mass is known to be associated with reduced strength, exercise tolerance, and functional autonomy. Conversely, basal metabolic rate showed a negative association with the “Can do, do do” group, possibly reflecting the influence of greater total body mass (particularly fat mass), which may increase resting energy expenditure but does not necessarily translate into better physical fitness^32^.

Diet quality is a recognized risk factor for COPD, influencing systemic inflammation and being associated with the intake of nutrients that support healthier body composition and higher physical activity levels. In our sample, nutritional intake did not differ significantly between groups. However, unfavorable body composition is well established in the literature as a predictor of lower physical performance and higher mortality in COPD, including variables such as FFM, SMM, and LBM^28, 33-37^. Our findings only partially reflect these associations, possibly because both groups had values within the normal range and due to the relatively small sample size in each group.

In the second analysis, only muscle-related bioimpedance variables and protein intake were included. A positive, although non-significant, trend was observed between higher protein intake, greater muscle mass, and the likelihood of belonging to the functionally active group (“Can do, do do”). This suggests that adequate protein intake may contribute to the maintenance of muscle mass and, consequently, better physical performance^38, 39^, even though this effect was not statistically confirmed—potentially due to sample size limitations and collinearity among muscle variables. Overall, the findings reinforce the importance of preserving and promoting muscle mass as a central component of maintaining functional capacity in patients with COPD.

According to Koolen’s classification, both of our groups have the expected level of physical capacity to perform PA; however, one group does not reach adequate PA levels. This raises an important question: if individuals are physically capable and have similar nutritional, biological, and clinical characteristics, why do some not engage in PA?

Socioeconomic and behavioral factors likely modulate PA levels in this population. Outdoor environments may pose barriers to PA due to traffic-related pollution. This challenge is heightened in Latin America, where higher pollution levels, warmer temperatures, and concerns regarding infrastructure safety may further hinder engagement in PA, even among those physically capable of performing it. Additionally, Latin American adults tend to engage more in transportation-based PA and less in leisure-based PA^40^. This may help explain our findings, as many participants with COPD were in an older age group and therefore no longer commuting for work.

Overall, we conclude that further studies are needed to investigate which additional factors influence PA in this population. A major limitation of our study was the small number of participants in the “Can’t do, don’t do” and “Can’t do, do do” quadrants, which prevented comparative analyses including all four quadrants. The inclusion of these groups would allow for a more comprehensive understanding of the model proposed by Koolen et al.

## CONCLUSION

Brazilians with COPD classified in quadrants I and II showed similar results in terms of body composition and food intake. A positive trend in skeletal muscle mass was observed for the ‘can do, do do’ group. These findings align with the pathophysiological model of COPD, in which the preservation of muscle mass and adequate protein intake support functional capacity and the maintenance of higher physical activity levels.

## Data Availability

Data corroborating the results of this study are available on request from the corresponding author [P.B].

## ACKNOWLEDGEMENTS

We would like to thank the São Paulo State University (UNESP) and the Laboratory for the Study of the Mucosecretory System (LEAMS), and the funding bodies, the National Council for Scientific and Technological Development (470742/2014-3) and the São Paulo Research Foundation (FAPESP for the funding; <u>#2017/10145-7</u>; #2018/04871-0; # 18/04870-3; #2017/10925-2).

## DECLARATION OF INTEREST

The authors of these studies have no conflicts of interest.

## AUTHORS CONTRIBUTIONS

Paloma Borges: conceptualization, analysis and data interpretation, writing, review and editing. Ana Paula Coelho Figueira Freire: conceptualization, analysis and data interpretation, review, editing and supervision. Fabiano Franscico de Lima: conceptualization, review and editing. Juliana Souza Uzeloto: data acquisition, review and editing. Bruna Spolador de Alencar Silva: data acquisition, writing, review and editing. Matheus Pedroso: data acquisition, analysis, review and editing. Luis Alberto Gobbo: data acquisition, review and editing. Isis Grigoletto: conceptualization, review and editing. Ercy Mara Cipulo Ramos: conceptualization, analysis and data interpretation, review, editing and supervision.

## REFERENCES

1. Xiang X, Huang L, Fang Y, Cai S, Zhang M. Physical activity and chronic obstructive pulmonary disease: a scoping review. BMC Pulm Med. 2022;22(1):301.

2. Silva EGd, Dourado VZ. Treinamento de força para pacientes com doença pulmonar obstrutiva crônica. Revista Brasileira de Medicina do Esporte. 2008;14.

3. Spruit MA, Augustin IM, Vanfleteren LE, Janssen DJ, Gaffron S, Pennings HJ, et al. Differential response to pulmonary rehabilitation in COPD: multidimensional profiling. The European respiratory journal. 2015;46(6):1625–35.

4. Cindy Ng LW, Mackney J, Jenkins S, Hill K. Does exercise training change physical activity in people with COPD? A systematic review and meta-analysis. Chron Respir Dis. 2012;9(1):17–26.

5. Spruit MA, Wouters EFM. Organizational aspects of pulmonary rehabilitation in chronic respiratory diseases. Respirology (Carlton, Vic). 2019;24(9):838–43.

6. Spruit MA, Pitta F, McAuley E, ZuWallack RL, Nici L. Pulmonary Rehabilitation and Physical Activity in Patients with Chronic Obstructive Pulmonary Disease. American journal of respiratory and critical care medicine. 2015;192(8):924–33.

7. Vogelmeier CF, Criner GJ, Martinez FJ, Anzueto A, Barnes PJ, Bourbeau J, et al. Global Strategy for the Diagnosis, Management, and Prevention of Chronic Obstructive Lung Disease 2017 Report. GOLD Executive Summary. American journal of respiratory and critical care medicine. 2017;195(5):557–82.

8. Freitas THPd, Souza DAFd. Corticosteróides sistêmicos na prática dermatológica. Parte I: Principais efeitos adversos. Anais Brasileiros de Dermatologia. 2007;82.

9. Dourado VZ, Godoy I. Recondicionamento muscular na DPOC: principais intervenções e novas tendências. Revista Brasileira de Medicina do Esporte. 2004;10.

10. Koolen EH, van Hees HW, van Lummel RC, Dekhuijzen R, Djamin RS, Spruit MA, et al. “Can do” versus “do do”: A Novel Concept to Better Understand Physical Functioning in Patients with Chronic Obstructive Pulmonary Disease. J Clin Med. 2019;8(3).

11. Vermeeren MAP, Creutzberg EC, Schols AMWJ, Postma DS, Pieters WR, Roldaan AC, et al. Prevalence of nutritional depletion in a large out-patient population of patients with COPD. Respiratory Medicine. 2006;100(8):1349–55.

12. Hogan D, Lan LT, Diep DT, Gallegos D, Collins PF. Nutritional status of Vietnamese outpatients with chronic obstructive pulmonary disease. Journal of human nutrition and dietetics : the official journal of the British Dietetic Association. 2017;30(1):83–9.

13. Rawal G, Yadav S. Nutrition in chronic obstructive pulmonary disease: A review. J Transl Int Med. 2015;3(4):151–4.

14. Ries AL, Kaplan RM, Limberg TM, Prewitt LM. Effects of pulmonary rehabilitation on physiologic and psychosocial outcomes in patients with chronic obstructive pulmonary disease. Ann Intern Med. 1995;122(11):823–32.

15. Choudhury G, Rabinovich R, MacNee W. Comorbidities and systemic effects of chronic obstructive pulmonary disease. Clin Chest Med. 2014;35(1):101–30.

16. Keogh E, Mark Williams E. Managing malnutrition in COPD: A review. Respir Med. 2021;176:106248.

17. Gómez-Martínez M, Rodríguez-García W, González-Islas D, Orea-Tejeda A, Keirns-Davis C, Salgado-Fernández F, et al. Impact of Body Composition and Sarcopenia on Mortality in Chronic Obstructive Pulmonary Disease Patients. Journal of Clinical Medicine. 2023;12(4):1321.

18. Ikeuchi T, Shingai K, Ichiki K, Jimi T, Kawano T, Kato K, et al. Effects of exercise intensity on nutritional status, body composition, and energy balance in patients with COPD: a randomized controlled trial. BMC Pulmonary Medicine. 2025;25(1):34.

19. O’Neill B, McKevitt A, Rafferty S, Bradley JM, Johnston D, Bradbury I, et al. A comparison of twice-versus once-weekly supervision during pulmonary rehabilitation in chronic obstructive pulmonary disease. Arch Phys Med Rehabil. 2007;88(2):167–72.

20. Costa TM, Costa FM, Moreira CA, Rabelo LM, Boguszewski CL, Borba VZ. Sarcopenia in COPD: relationship with COPD severity and prognosis. Jornal brasileiro de pneumologia : publicacao oficial da Sociedade Brasileira de Pneumologia e Tisilogia. 2015;41(5):415–21.

21. Miller MR, Hankinson J, Brusasco V, Burgos F, Casaburi R, Coates A, et al. Standardisation of spirometry. The European respiratory journal. 2005;26(2):319–38.

22. Neder JA, Andreoni S, Castelo-Filho A, Nery LE. Reference values for lung function tests. I. Static volumes. Braz J Med Biol Res. 1999;32(6):703–17.

23. Fisberg RM, Marchioni DML, Colucci-Paternez ACA. Avaliação do consumo alimentar e da ingestão de nutrientes na prática clÍnica. Arquivos Brasileiros de Endocrinologia & Metabologia. 2009;53(5):617–24.

24. Slater B, Philippi ST, Fisberg RM, Latorre MR. Validation of a semi-quantitative adolescent food frequency questionnaire applied at a public school in São Paulo, Brazil. Eur J Clin Nutr. 2003;57(5):629–35.

25. Troosters T, Gosselink R, Decramer M. Six minute walking distance in healthy elderly subjects. The European respiratory journal. 1999;14(2):270–4.

26. Holland AE, Spruit MA, Troosters T, Puhan MA, Pepin V, Saey D, et al. An official European Respiratory Society/American Thoracic Society technical standard: field walking tests in chronic respiratory disease. The European respiratory journal. 2014;44(6):1428–46.

27. Van Remoortel H, Raste Y, Louvaris Z, Giavedoni S, Burtin C, Langer D, et al. Validity of six activity monitors in chronic obstructive pulmonary disease: a comparison with indirect calorimetry. PloS one. 2012;7(6):e39198.

28. Schols AM, Mostert R, Soeters PB, Wouters EF. Body composition and exercise performance in patients with chronic obstructive pulmonary disease. Thorax. 1991;46(10):695–9.

29. Ricieri DdV. Analysis of body impedance in COPD patients for functional kinesiologic diagnostic. Fisioterapia Brasil. 4:85–93.

30. Jaitovich A, Barreiro E. Skeletal Muscle Dysfunction in Chronic Obstructive Pulmonary Disease. What We Know and Can Do for Our Patients. American journal of respiratory and critical care medicine. 2018;198(2):175–86.

31. Li P, Li J, Wang Y, Xia J, Liu X. Effects of Exercise Intervention on Peripheral Skeletal Muscle in Stable Patients With COPD: A Systematic Review and Meta-Analysis. Frontiers in medicine. 2021;8:766841.

32. Lührmann PM, Herbert BM, Neuhäuser-Berthold M. Effects of fat mass and body fat distribution on resting metabolic rate in the elderly. Metabolism: clinical and experimental. 2001;50(8):972–5.

33. Ricieri D. Análise da bioimpedância corporal em portadores de DPOC: uma visão para o diagnóstico cinesiológico funcional. Fisioterapia Brasil. 2009;4:85.

34. Eisner MD, Blanc PD, Sidney S, Yelin EH, Lathon PV, Katz PP, et al. Body composition and functional limitation in COPD. Respiratory research. 2007;8(1):7.

35. Vestbo J, Prescott E, Almdal T, Dahl M, Nordestgaard BG, Andersen T, et al. Body mass, fat-free body mass, and prognosis in patients with chronic obstructive pulmonary disease from a random population sample: findings from the Copenhagen City Heart Study. American journal of respiratory and critical care medicine. 2006;173(1):79–83.

36. Gologanu D, Ionita D, Gartonea T, Stanescu C, Bogdan MA. Body composition in patients with chronic obstructive pulmonary disease. Maedica. 2014;9(1):25–32.

37. Pitta F, Troosters T, Spruit MA, Probst VS, Decramer M, Gosselink R. Characteristics of physical activities in daily life in chronic obstructive pulmonary disease. American journal of respiratory and critical care medicine. 2005;171(9):972–7.

38. Bernardes S, Eckert IDC, Burgel CF, Teixeira PJZ, Silva FM. Increased energy and/or protein intake improves anthropometry and muscle strength in chronic obstructive pulmonary disease patients: a systematic review with meta-analysis on randomised controlled clinical trials. The British journal of nutrition. 2022:1–18.

39. Ahmadi A, Eftekhari MH, Mazloom Z, Masoompour M, Fararooei M, Eskandari MH, et al. Fortified whey beverage for improving muscle mass in chronic obstructive pulmonary disease: a single-blind, randomized clinical trial. Respiratory research. 2020;21(1):216.

40. Jáuregui A, Salvo D, Medina C, Barquera S, Hammond D. Understanding the contribution of public- and restricted-access places to overall and domain-specific physical activity among Mexican adults: A cross-sectional study. PloS one. 2020;15(2):e0228491.

